# People’s expectations about the likely duration of common acute infections: a national survey of the Australian public

**DOI:** 10.1101/2024.06.22.24309159

**Authors:** Kwame Peprah Boaitey, Mina Bakhit, Mark Jones, Tammy C Hoffmann

## Abstract

**Objectives:** To explore people’s expectations about the likely duration of acute infections that are commonly managed in primary care and if care is sought for these infections, reasons for doing so.

**Design:** A cross-sectional online survey.

**Participants:** A nationwide sample of 589 Australian residents, ≥18 years old with representative quotas for age and gender, recruited via an online panel provider.

**Outcome measures:** For eight acute infections, participants’ estimated duration of each, time until they would seek care, and reasons for seeking care.

**Results:** For four infections, participants’ mean estimates of duration were within an evidence-based range - common cold (7.2 days), sore throat (5.2 days), acute otitis media (6.2 days), and impetigo (8.3 days); and >70% of participants estimated a duration within the range. However, participants estimated mean duration was shorter than evidence-based estimates for acute cough (7.6 days), sinusitis (5.6 days), conjunctivitis (5.7 days), and uncomplicated urinary tract infections (5.4 days); and >60% of participants underestimated the duration. Of the 589 participants, 365 (62%) indicated they were unlikely to routinely seek care for self-limiting infections. Most common reasons for care-seeking were severe or worsening symptoms, a desire for quick recovery, and fear of progression to complications. After being shown typical durations, the proportion of participants who reported having no concerns waiting for spontaneous resolution while managing symptoms with over-the-counter medications ranged across the infections and was highest for common cold (68%) and lowest for UTI (31%).

**Conclusion:** Participants underestimated the duration of some infections compared to evidence-based estimates and were accurate in their estimates for other infections. Many stated that they would not be concerned about waiting for illnesses to self-resolve after learning the typical duration. Communicating the expected duration of common acute infections during routine consultations can help manage patients’ expectations of recovery and need to seek care.

**Strengths and limitations of this study**

- Representative sample with quotas for age and sex.
- First Australian survey of people’s expected duration of common acute infections.
- Participants responded to hypothetical scenarios and did not have the infections.
- This was a convenience sample from one online panel provider and not a true random sample.

## INTRODUCTION

The overuse of antibiotics is a contributor to the global health threat of antibiotic resistance.^1,2^ Despite evidence that antibiotics provide limited benefit and no-to-minimal reduction in the duration of many common acute infections,^3,4^ they continue to be frequently prescribed. ^5,6^ In primary care, antibiotics are often unnecessarily prescribed for acute infections, such as acute respiratory infections (ARIs), uncomplicated urinary tract infections (UTIs) and some skin and soft tissue infections (SSTIs).^7,8^ For example, in 2019, 82% of Australians who presented at primary care with acute bronchitis were prescribed antibiotics.^9^

There are various reasons why antibiotics are unnecessarily prescribed, including diagnostic uncertainty, concerns about damaging patient-clinician relationships, suboptimal communication between patients and clinicians, and articulated or perceived patient expectations for antibiotics.^10-12^ Effective communication between patients and clinicians about the expected duration of illnesses and how much it might or might not be reduced by antibiotics may help to set realistic expectations about the recovery timeframe for patients and help counter patients’ misperceptions about the need for antibiotics.^13,14^

Central to managing expectations is knowing about the natural history of the illness and for acute illnesses, particularly its typical duration. Natural history can be defined as the course of a disease process over time in the absence of treatment.^15^ A study of adults’ expectation of acute cough duration found that participants predicted a mean duration of 7.2 to 9.3 days, compared with published literature estimates of 17.8 days.^16^ This discrepancy leads individuals to seek care and expect antibiotics if they believe their infection has persisted longer than they thought it should.^17^ Beyond this study, there has been little exploration of individuals’ knowledge about the duration of common acute infections. In a sample of Australian adults, this study aimed to determine people’s expectations about the duration of common acute infections and explore reasons for deciding to seek care.

## METHODS

### Study design and participants

A cross-sectional, online survey of Australian residents was conducted from 10^th^ to 17^th^ February 2024. To be eligible, participants had to be an Australian resident ≥18 years old, not a health professional or health professional student, and able to read and understand English. We excluded health professionals as their knowledge about common infections may differ from the general population.

### Recruitment and study procedure

A national sample with representation quotas for age and gender was recruited through an online independent panel provider, Dynata (https://www.dynata.com/), which specialises in using algorithm-based sampling tools for registered members who have previously consented to participate in online surveys. At survey commencement, potential participants received an information form explaining the study’s aims and their right to withdraw at any time. Participants’ continuation of the survey was accepted as informed consent. To ensure the validity and uniqueness of responses, Dynata used a captcha at the commencement of each survey and an IP-digital stamp for each participant. Participants were compensated based on Dynata’s pre-agreed structured incentive scheme policy, which allowed participants to redeem from a range of gift cards, points programs, charitable contributions and partner products or services. An invitation to the study was emailed to participants individually using an automated router. See Supplementary File 1 for more information on Dynata Australia’s demographics, sampling, and recruitment procedures.

### Patient and public involvement

We piloted the survey face-to-face with a convenience sample of eligible participants (n=10) to establish face and content validity. Feedback from participants was used to refine some questions to ensure their clarity. To test for technical issues, we piloted the survey online with 10% of the sample size needed for the study. Participants recruited for the pilot were not invited to participate in the subsequent survey and the pilot data were not included in the analysis.

### Data collection and outcome measurement

The survey focussed on common infections that are typically managed in primary care and often self-resolve without treatment, other than for managing symptoms such as fever. We asked about these eight infections: acute cough (acute bronchitis), common cold, sore throat, middle ear infection (acute otitis media), sinusitis, viral conjunctivitis, uncomplicated urinary tract infection (UTI), and school sores (non-bullous impetigo). The survey (Supplementary File 2) contained three sections: A) 12 demographic questions and a single question about preferences for passive or aggressive health treatment (Medical Maximiser or Minimiser Scale); ^18^ B) three questions about seeking care for common infections in general and possible influences; and C) for each infection, four questions about the infection’s estimated duration, waiting time until seeking care, and concerns about waiting for infections to run its course. We randomised the order in which the infections were presented to minimise order bias. We did not allow participants to revise their responses to questions that they had already completed within the survey.

### Sample size

The sample size needed was calculated based on the accuracy of the estimate of the proportion of the eligible population who decided to seek care. Assuming a margin of error of 5% and a 95% confidence level, we required a minimum of 426 participants. This calculation allowed for up to 10% missing data on the decision to seek care question.

### Data analysis

Data were analysed descriptively with Stata/MP 16.1. For each infection, we calculated the mean duration of infections as well as the proportion of participants whose estimated duration was within the evidenced-based range or an over- or under-estimation. Estimates were deemed “correct” if they were +/- 1 day of the evidence-based estimates obtained from systematic reviews (references are in Figure 1). Participants’ responses to likelihood to seek care questions (Supplementary File 2, question 14) were collapsed into three categories: “Unlikely” (combining “Very Unlikely” and “Unlikely”), “Neutral”, and “Likely” (combining “Likely” and “Very Likely”). Responses to open-ended questions (Supplementary File 2, questions 15, 16, A3 - H3) were independently coded and grouped into common categories by two authors, using Microsoft® Excel® 365 (Microsoft, Redmond, WA, USA). To analyse the ranking data (Supplementary File 2, questions A4-H4), we assigned a score based on each rank (i.e., 5 for 1^st^, 4 for 2^nd^, and so on, excluding the rank for “other”). The scores were summed for each reason for all respondents. We calculated the average score for each reason to determine the overall importance assigned by respondents.

**Figure 1:**
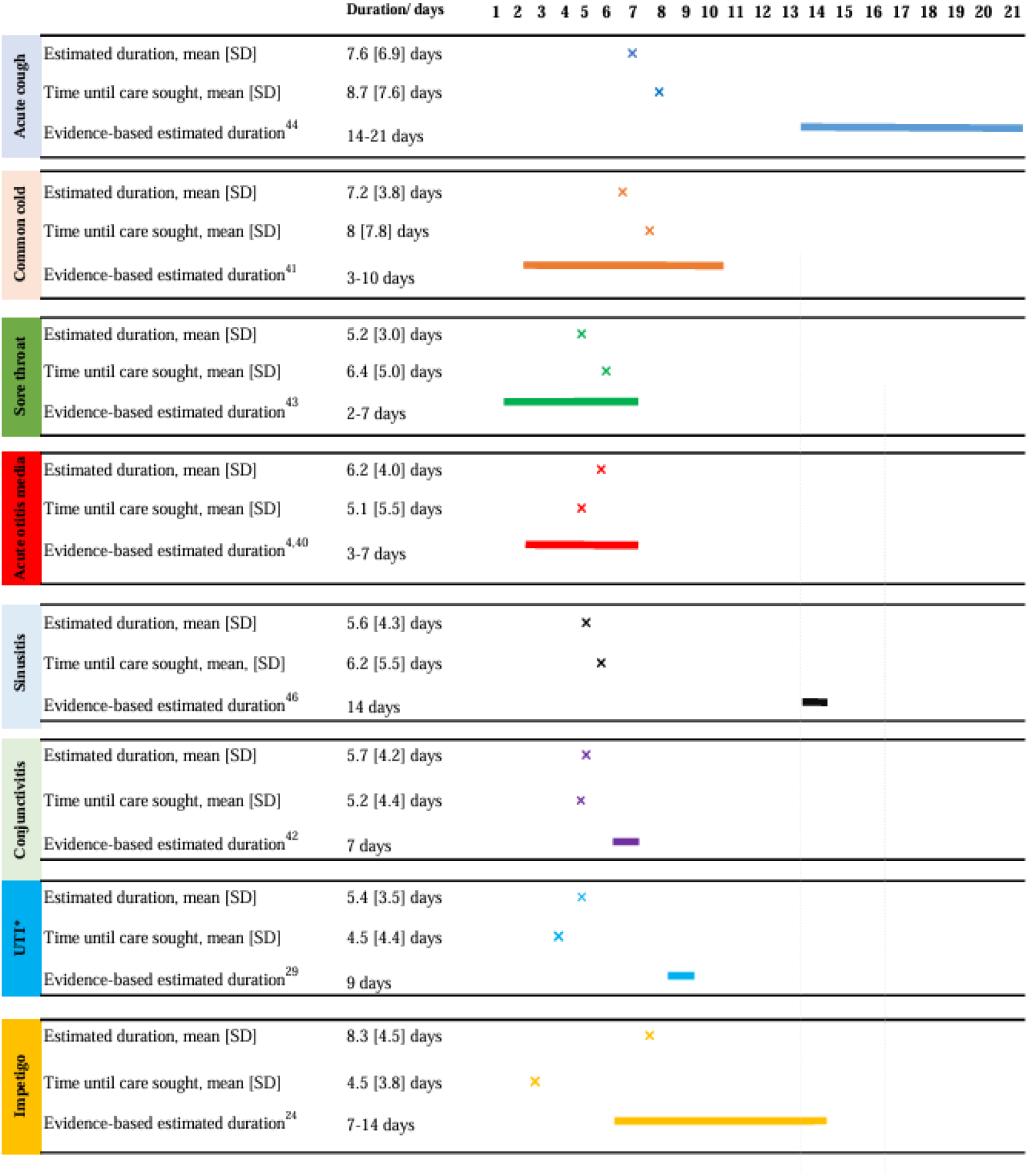
Participants’ estimated mean duration of acute infections and time until seeking care, and evidence-based estimates of duration (evidence sources ^4,24,29,40-45^). **SD:** standard deviation, \***UTI**: uncomplicated Urinary Tract Infections.

### Ethical approval

The study was approved by the Bond University Human Research Ethics Committee (reference number: KB03170)

## RESULTS

### Participant characteristics

Of the 1288 potential participants who completed the survey, 699 responses were screened as ineligible, leaving responses from 589 respondents for analysis (see Supplementary File 3 for participant flow chart with reasons for ineligibility).

Participants’ characteristics are reported in Table 1. Just over half (n=348, 59%) were female, 42% (n=247) were within the age category of ≥56 years, and 29% (n=170) had non-adult children living with them. Just over half (n=346, 59%) indicated a preference for a more passive approach to healthcare, and 62% (n=366) indicated that, in general, they were unlikely to seek healthcare for a self-limiting acute infection.

**Table 1.**
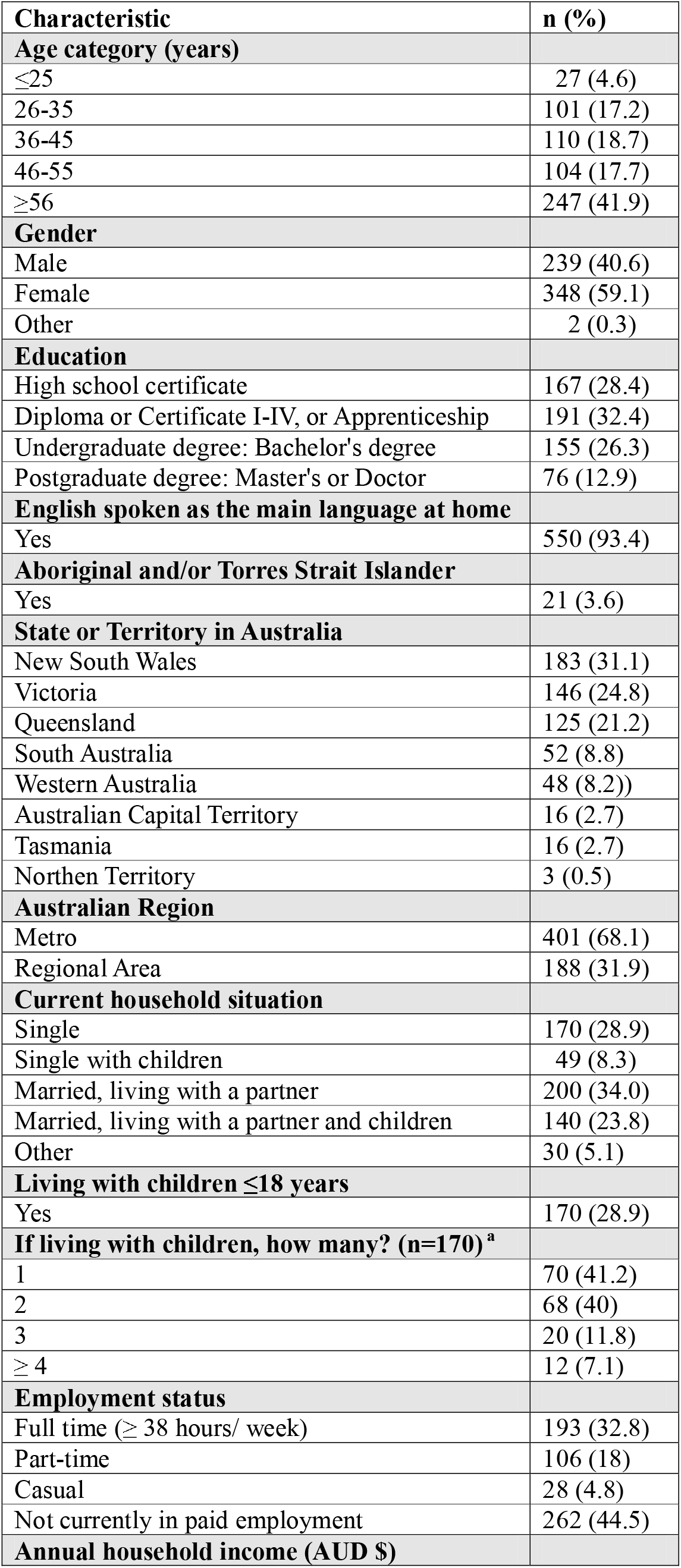
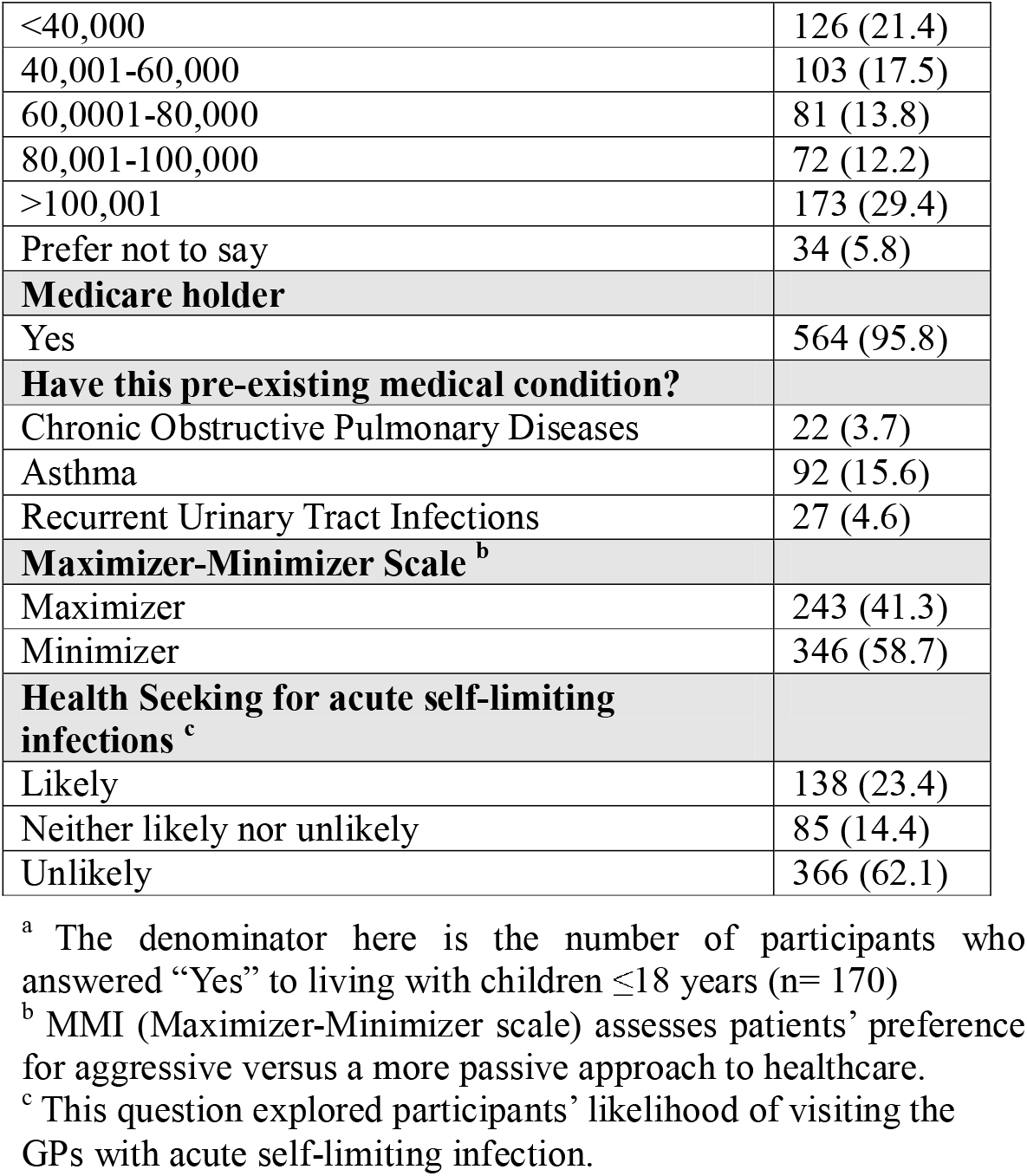
Participant characteristics (n=589, unless stated otherwise)

### Estimated duration of infections and time until seeking care

Figure 1 shows participants estimated mean duration for each infection, estimated mean time until seeking care, and an evidence-based estimate of the duration. For cough, sinusitis, conjunctivitis and uncomplicated UTI, participants’ mean estimated duration was shorter than the evidenced-based estimates. For common cold, acute otitis media, sore throat, and non-bullous impetigo, participants’ mean estimated duration were within the evidence-based range. When participants’ responses were analysed as the proportion whose duration estimate was correct, or an over- or under-estimate (Fig 2), the majority (≥50%) of participants underestimated the duration of cough (83%), sinusitis (92%), conjunctivitis (63%), and uncomplicated UTI (88%), compared with evidence-based estimates. Whereas the majority (≥50%) of participants provided a correct duration estimate for sore throat (91%), common cold (86%), acute otitis media (78%), and impetigo (65%).

**Figure 2:**
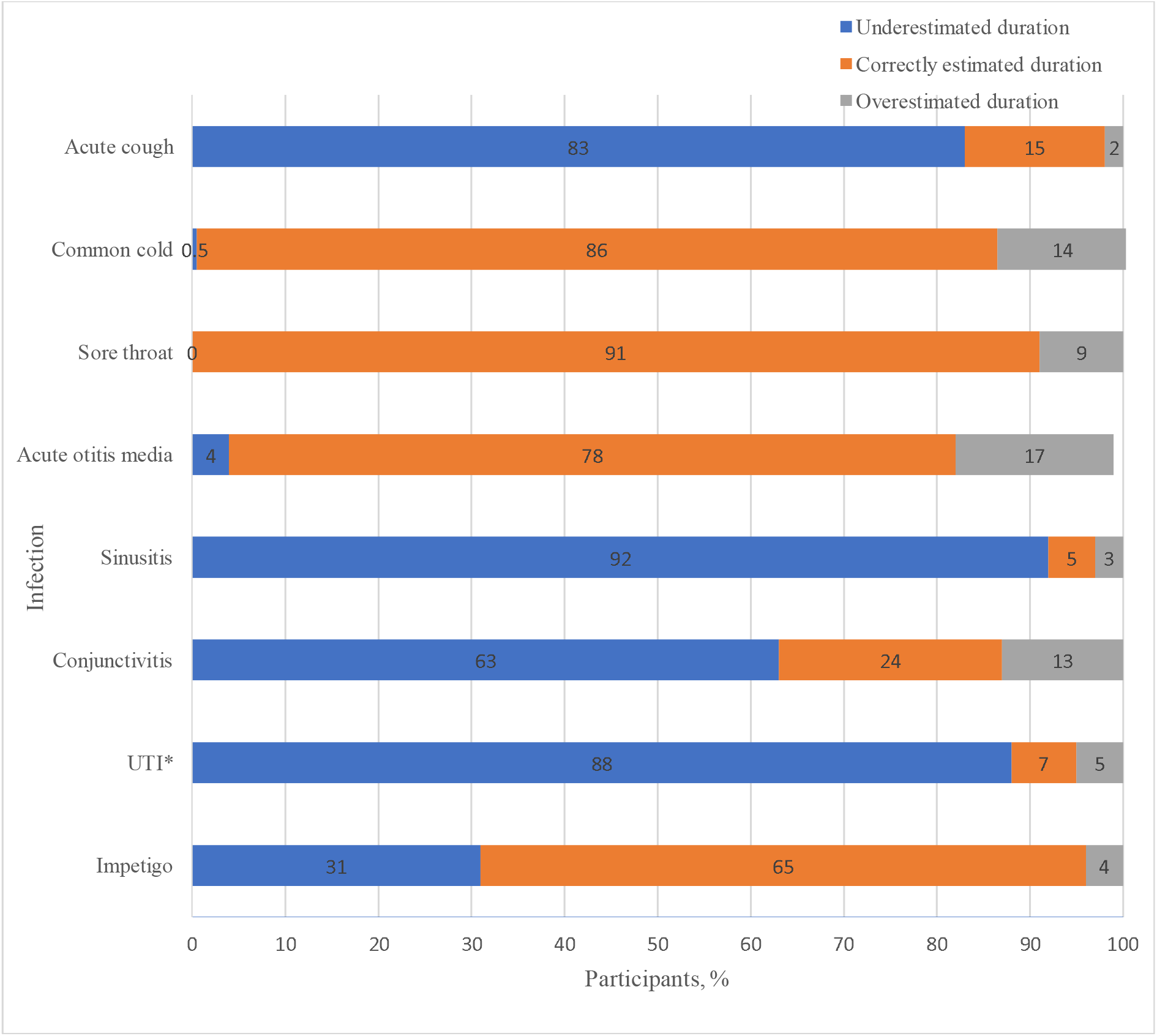
Percentage of participants predicting a correct estimate, underestimate, or overestimate of the duration of each of the acute infections. **UTI**: uncomplicated Urinary Tract Infections

The mean amount of time that participants reported they would wait before seeking care was within +/-1 day of their estimated duration for all infections (Fig 1), with one exception. For impetigo, participants indicated they would seek care, on average, 4 days earlier than when they estimated it would have resolved by.

### Reasons for seeking care

Figure 3 presents participants’ reasons for seeking care, for each infection, based on the response options provided in the survey. The ranked response pattern was similar across most infections, with approximately an equal number of responses across the five response options provided to participants. An exception was acute cough, where participants ranked “wanting to reduce the impact on daily life” as the most important reason for seeking care, followed by “symptoms taking too long to get better”. For common cold, sore throat and sinusitis, participants ranked “wanting to get better faster” as their main reason for seeking care, whereas for acute otitis media, uncomplicated UTI, and impetigo, the main reason was “wanting to prevent complications.”

**Figure 3:**
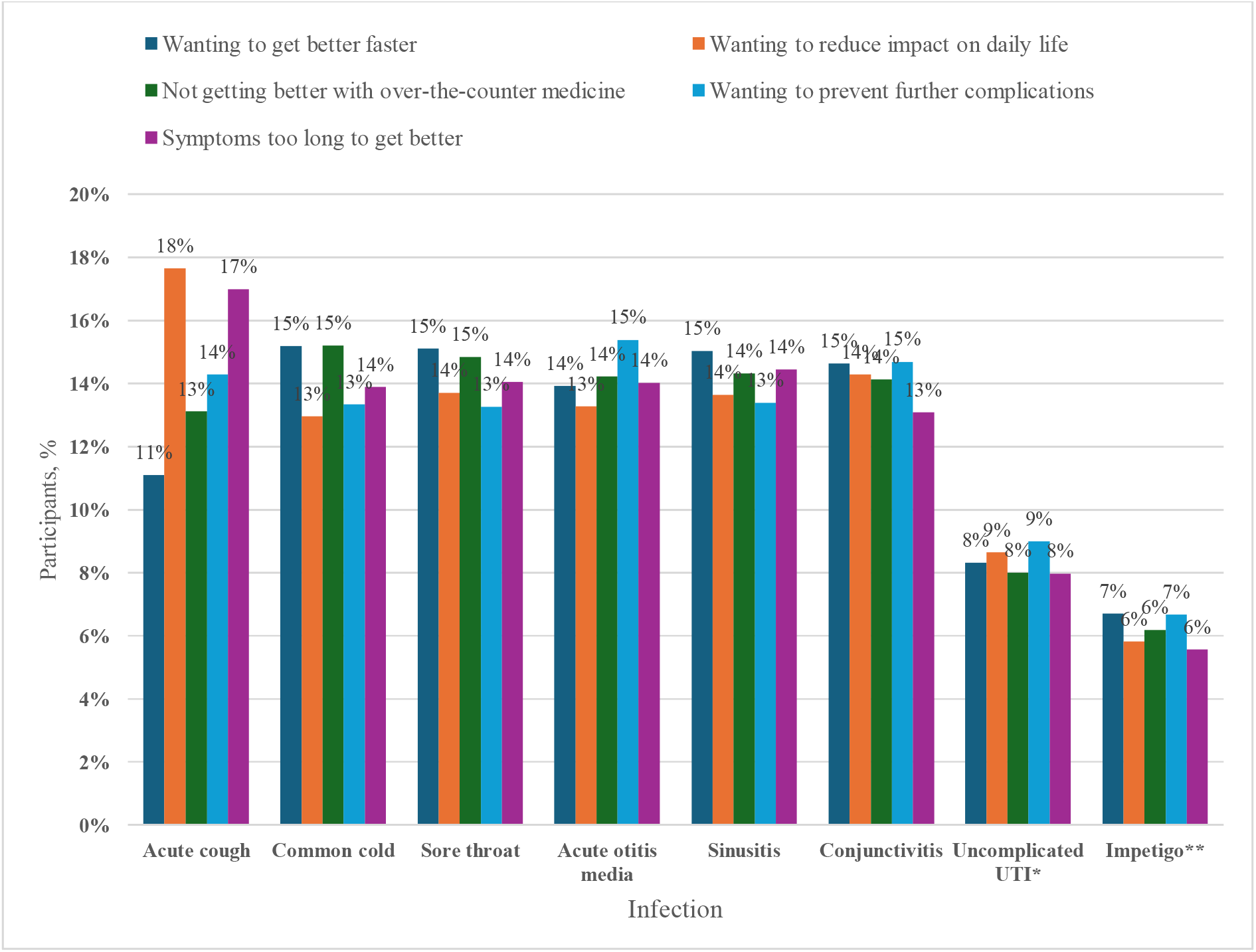
Participants’ reasons for seeking care, ranked, for each infection. ^*^Urinary tract infection (applies to only female respondents n=348) ^**^only answered by respondents who answered “yes” to knowledge about uncomplicated impetigo (n=170)

An analysis of the free-text responses to the question about reasons for seeking care in general for an acute infection identified similar categories among participants. The most frequently reported reasons for seeking care were a desire to recover quickly or experiencing severe, concerning, or unfamiliar symptoms. However, many participants reported that when it was their child that had an infection, t they were more likely to seek care early, regardless of its severity, nature, or duration.

After the survey questions revealed to participants the evidence-based duration estimate of each infection, participants were asked if they had concerns about waiting this length of time to seek care and if so, to list their concern/s. The proportion who had no concerns about waiting for the infection to resolve on its own and not seek care beyond using over-the-counter medicines varied across the infections. It was highest for common cold (68%), sore throat (59%) and impetigo (51%), followed by conjunctivitis (47%), cough (46%), sinusitis (44%), acute otitis media (36%), and UTI (31%). The most commonly reported concerns about waiting to seek care for each infection were similar to those mentioned before the evidence-based duration estimate was shown. One notable difference was that now, the least commonly indicated concern for care-seeking was that symptoms were taking too long to improve. Other factors mentioned in free-text comments about influences on care-seeking behaviours were the ease of access to healthcare services and the associated costs.

## DISCUSSION

This survey explored people’s estimates of the duration of common acute infections and reasons that influence their decision to seek care. The majority of participants underestimated the duration of cough, sinusitis, conjunctivitis, and uncomplicated UTI, whereas duration estimates were within the evidence-based range for common cold, acute otitis media, sore throat, and non-bullous impetigo. The most frequent reasons for seeking care were when symptoms perceived as severe, a desire to recover faster, and being worried about possible complications if left untreated.

When an illness persists longer than they expect it to, people may seek care and may request or expect antibiotics, believing antibiotics will accelerate their recovery.^10,17^ For four of the infections explored in our study (cough, sinusitis, conjunctivitis, UTI), we found that participants underestimated their duration. We are aware of only two studies that have quantitatively explored people’s expectations of the duration of common infections. In a study of American adults’ expectations of cough duration, participants underestimated the duration of cough by about 8 days, ^16^ which was similar to our finding. A study conducted in Hong Kong on people’s expectations of the duration of an upper respiratory tract infection found that participants anticipated their infection to last for an average of 7.4 days less than the evidence-based estimate. ^19^ People may underestimate the duration of infections because of low awareness of the typical course of the infection, previous experiences, and a common misperception that antibiotics are necessary to treat the infection and can reduce its duration. ^10,16,20^ Additionally, public health campaigns about self-limiting infections have largely not focussed on infections such as cough, sinusitis, conjunctivitis, and UTI.

Participants’ expectations about the duration of some infections (common cold, sore throat, acute otitis media, impetigo) were within evidence-based estimates. One possible reason, for at least the three acute respiratory infections, is that various public campaigns in Australia (such as by the National Prescribing Service ^21^ and as part of Choosing Wisely ^22^) have communicated that most acute respiratory infections do not need antibiotics and they get better on their own. It is not clear why participants’ estimates of impetigo duration were within the evidence-based range. It may partly be because the range is large as there are far fewer studies of the natural history of impetigo than there are of acute respiratory infections. ^23,24^ Impetigo was also the one condition where the mean number of days that participants indicated they would wait before seeking care was not within one day of the mean estimated infection duration. Participants indicated they would seek care, on average, 4 days earlier, with some participants providing reasons for this as concern about the risk of scarring, impact on daily life, and wanting to avoid spreading the infections.

Just over half of our participants indicated that they would usually not seek care for common acute self-limiting infections, which is consistent with the findings of previous research. ^17,25^ In a qualitative study that explored participants’ decisions to attend their family physician in Canada, participants indicated they do not routinely seek care for acute respiratory tract infections. ^17^ Similarly, a study in the United Kingdom that explored women’s journey from self-care to GP care when they had UTI symptoms found that most participants do not routinely consult their GPs for UTI and often initiate self-care, followed by a period of monitoring and only consult when they thought self-care had failed.^25^

We found that the main reasons people gave for seeking care were worsening symptoms, severe symptoms, a desire to get better quickly and to reduce the risk of complications. This is similar to the findings in previous studies.^17,25,26^ In a qualitative study exploring triggers of care-seeking for women with UTI, failure of symptoms to alleviate, symptom duration and escalation, and concern about illness seriousness were the major drivers of primary care visits.^25^ Concern about the risk of complications by not treating is a key contributor to patients’ expectations of antibiotics, which can influence prescribing decisions. ^10,11,27^ This is despite research showing that, for most of the infections studied, the risk of complications without antibiotic use is low. ^28-30^

Understanding why people seek care and what they expect from the visit can be facilitated if clinicians actively elicit and discuss expectations during consultations.^31^ This can enable clinicians to provide reassurance, address any misperceptions, explain the options and provide information about symptoms to monitor or reasons for when they should reconsult. ^31^ Consultations for acute infections are well suited to shared decision making where the options of taking or not taking antibiotics can be discussed, along with the benefits and harms of each option. ^32,33^ Along with shared decision making, clinicians can also use other antimicrobial stewardship activities such as delayed prescribing. ^34^ Integral to both strategies is knowing the evidenced-based estimates of infection duration to guide the discussion.

Participants in our study were least concerned about needing to seek care and the time taken for symptoms to improve after they were presented with the typical duration of the infections. Similarly, a study in the United Kingdom that explored patients’ understanding and management of conjunctivitis found that patients who learnt about the typical duration of conjunctivitis were more likely to self-manage and less likely to visit their clinician.^35^ A recent qualitative study of Australian GPs showed that GPs found natural history information such as duration valuable when discussing antibiotic use for self-limiting conditions.^36^ However, GPs do not consistently discuss the likely duration of infections during consultations ^14^ because such information is not routinely available in clinical resources such as clinical practice guidelines. ^37^

Ways of providing natural history information to clinicians so that it can be incorporated into consultations should be explored in future research. Beyond GP consultations, future research could also consider how to increase public awareness of the typical duration of the infections for which duration was underestimated (e.g. cough). This may include public awareness campaigns or information provided via community pharmacies as that is where many patients visit as their first point of care. ^38^

### Strengths and limitations

A strength of the study is the sample’s representativeness of the Australian adult population in terms of age, sex, and location.^39^ However, there are several limitations. Firstly, participants were more highly educated than the general Australian adult population and were recruited from one panel provider, which may affect the generalisability of the findings and introduce selection bias. Secondly, participants in our study responded to hypothetical scenarios of acute infections, and their actual behaviour may differ if or when they had these infections. Thirdly, each acute respiratory infection was considered separately, even though some symptoms, such as sore throat, can occur in other infections. This overlap may influence participants’ health-seeking behaviour. Finally, the evidence-based infection duration ranges are only approximations and depend on the number and quality of existing studies. Although the estimates were based on a recent scoping review that mapped natural history evidence, ^40^ as well as a systematic review that examined natural history information inclusion in clinical practice guidelines ^37^ for some infections (such as UTI and impetigo), the available evidence is limited, with wide ranges and some uncertainty about infection duration.

## Conclusion

Our study found that for some acute infections, people underestimate the likely duration. This may contribute to people seeking care and possibly antibiotics. For some infections, people’s estimates were within an evidence-based range. The study highlights the complex interplay of individual concerns that influence care-seeking behaviour for common acute infections. As many participants generally felt comfortable self-managing their infections after being informed about the likely evidence-based duration, such information should be available across primary care settings, including pharmacies and general practices, to enhance patients’ understanding and self-management of common infections.

X: Kwame Peprah Boaitey @PeprahBk, Mina Bakhit @Mina_Bakhit, Tammy Hoffmann @Tammy_Hoffmann

## Data Availability

All data produced in the present work are contained in the manuscript

## Author contributions

KPB, MB, and TH conceived the study. All authors contributed equally to the study design. KPB drafted the initial survey with critical inputs from MB and TH. Data collection was managed by Dynata under the direction of KPB. Data were analysed by KPB, MJ and MB with inputs from TH. KPB created the figures, tables, and drafted the manuscript with consultation from MB, MJ, and TH. The authors reviewed the manuscript and approved the final version for submission. KPB is responsible for the overall content as the guarantor.

## Patient and public involvement

Members of the public were involved in the piloting of this research. Refer to the Methods section for further details.

## Patient consent for publication

Not required.

Ethics approval Bond University Human Research Ethics Committee provided ethics approval (number: KB03170).

## Provenance and peer review

Not commissioned, externally peer reviewed.

## Data availability statement

All data relevant to this study are available in the article or uploaded as supplementary information.

## Supplemental material

This content has been supplied by the author(s). It has not been vetted by BMJ Publishing Group Limited (BMJ) and may not have been peer-reviewed. Any opinions or recommendations discussed are sorely those of the author(s) and are not endorsed by BMJ. BMJ disclaims all liability and responsibility arising from any reliance placed on the content. Where the content includes any translated material, BMJ does not warrant the accuracy and reliability of the translations (including but not limited to local regulations, clinical guidelines, terminology, drug names and drug dosages), and is not responsible for any error and/or omission arising from translation and adaptation or otherwise.

**Open access**

## References

1. Murray CJL, Ikuta KS, Sharara F, et al. Global burden of bacterial antimicrobial resistance in 2019: a systematic analysis. The Lancet. 2022;399(10325):629–655. doi:10.1016/S0140-6736(21)02724-0

2. World Health Organisation. Global action plan on antimicrobial resistance. 2015. 9789241509763. Accessed December 06. https://www.who.int/publications/i/item/9789241509763

3. Spinks A, Glasziou PP, Del Mar CB. Antibiotics for sore throat. Cochrane Database Syst Rev. 2013;(11)doi:10.1002/14651858.CD000023.pub4

4. Venekamp RP, Sanders SL, Glasziou PP, Del Mar CB, Rovers MM. Antibiotics for acute otitis media in children. Cochrane Database Syst Rev. 2015;(6)doi:10.1002/14651858.CD000219.pub4

5. Bell BG, Schellevis F, Stobberingh E, Goossens H, Pringle M. A systematic review and meta-analysis of the effects of antibiotic consumption on antibiotic resistance. BMC Infect Dis. 2014/01/09 2014;14(1):13. doi:10.1186/1471-2334-14-13

6. McCullough AR, Pollack AJ, Hansen MP, et al. Antibiotics for acute respiratory infections in general practice: comparison of prescribing rates with guideline recommendations. Medical Journal of Australia. 2017;207(2):65–69.

7. Pouwels KB, Dolk FCK, Smith DRM, Robotham JV, Smieszek T. Actual versus ‘ideal’ antibiotic prescribing for common conditions in English primary care. Journal of Antimicrobial Chemotherapy. 2018;73(Suppl_2):19–26. doi:10.1093/jac/dkx502

8. Sulis G, Daniels B, Kwan A, et al. Antibiotic overuse in the primary health care setting: a secondary data analysis of standardised patient studies from India, China and Kenya. BMJ Global Health. Sep 2020;5(9)doi:10.1136/bmjgh-2020-003393

9. Australian Commission on Safety and Quality in Health Care. AURA 2023: Fifth Australian report on antimicrobial use and resistance in human health. 2023:84. Accessed December 6. https://www.safetyandquality.gov.au/our-work/antimicrobial-resistance/antimicrobial-use-and-resistance-australia-aura/aura-2023-fifth-australian-report-antimicrobial-use-and-resistance-human-health

10. Coxeter PD, Mar CD, Hoffmann TC. Parents’ expectations and experiences of antibiotics for acute respiratory infections in primary care. Annals of Family Medicine. 2017;15(2):149–154.

11. Lucas PJ, Cabral C, Hay AD, Horwood J. A systematic review of parent and clinician views and perceptions that influence prescribing decisions in relation to acute childhood infections in primary care. Scandinavian Journal of Primary Health Care. 2015/01/02 2015;33(1):11–20. doi:10.3109/02813432.2015.1001942

12. Teixeira Rodrigues A, Roque F, Falcão A, Figueiras A, Herdeiro MT. Understanding physician antibiotic prescribing behaviour: a systematic review of qualitative studies. International Journal of Antimicrobial Agents. 2013;41(3):203–212.

13. Del Mar C, Hoffmann T, Bakhit M. How can general practitioners reduce antibiotic prescribing in collaboration with their patients? Australian Journal of General Practice. Jan-Feb 2022;51(1-2):25–30. doi:10.31128/ajgp-07-21-6084

14. Butler CC, Rollnick S, Kinnersley P, Tapper-Jones L, Houston H. Communicating about expected course and re-consultation for respiratory tract infections in children: an exploratory study. British Journal of Geneberal Practice. 2004;54(504):536–538.

15. Jose L T. A narrative review of natural history of diseases and continuity of care in family medicine. Archives of Community Medicine and Public Health. 2017:41–47. doi:10.17352/2455-5479.000023

16. Ebell MH, Lundgren J, Youngpairoj S. How long does a cough last? Comparing patients’ expectations with data from a systematic review of the literature. Annals of Family Medicine. 2013;11(1):5–13. doi:10.1370/afm.1430

17. Mortazhejri S, Patey AM, Stacey D, Bhatia RS, Abdulla A, Grimshaw JM. Understanding determinants of patients’ decisions to attend their family physician and to take antibiotics for upper respiratory tract infections: a qualitative descriptive study. BMC Family Practice. 2020/06/24 2020;21(1):119. doi:10.1186/s12875-020-01196-9

18. Scherer LD, Zikmund-Fisher BJ. Eliciting medical maximizing-minimizing preferences with a single question: development and validation of the MM1. Medical Decision Making. 2020;40(4):545–550. doi:10.1177/0272989×20927700

19. Lam TP, Lam KF, Wun YT, Sun KS. How long do the Hong Kong Chinese expect their URTI to last? – effects on antibiotic use. BMC Pulmunary Medicine. 2015/03/15 2015;15(1):23. doi:10.1186/s12890-015-0018-y

20. Boiko O, Gulliford MC, Burgess C. Revisiting patient expectations and experiences of antibiotics in an era of antimicrobial resistance: qualitative study. Health Expectation. Oct 2020;23(5):1250–1258. doi:10.1111/hex.13102

21. Wutzke SE, Artist MA, Kehoe LA, Fletcher M, Mackson JM, Weekes LM. Evaluation of a national programme to reduce inappropriate use of antibiotics for upper respiratory tract infections: effects on consumer awareness, beliefs, attitudes and behaviour in Australia. Health promotion international. Mar 2007;22(1):53–64. doi:10.1093/heapro/dal034

22. Muscat DM, Thompson R, Cvejic E, et al. Randomized trial of the Choosing Wisely consumer questions and a shared decision-making video intervention on decision-making outcomes. Medical Decision Making. 2023/08/01 2023;43(6):642–655. doi:10.1177/0272989X231184461

23. Bakhit M, Hoffmann T, Santer M, et al. Comparing the quantity and quality of randomised placebo-controlled trials of antibiotics for acute respiratory, urinary, and skin and soft tissue infections: a scoping review. British Journal of General Practice Open. 2020;4(4)

24. Hoffmann TC, Peiris R, Glasziou P, Cleo G, Mar CD. Natural history of non-bullous impetigo: a systematic review of time to resolution or improvement without antibiotic treatment. British Journal of General Practice. 2021;71(704):e237–e242.

25. Leydon GM, Turner S, Smith H, Little P. The journey from self-care to GP care: a qualitative interview study of women presenting with symptoms of urinary tract infection. British Journal of General Practice. Jul 2009;59(564):e219–25. doi:10.3399/bjgp09X453459

26. van der Velden AW, Sessa A, Altiner A, Pignatari ACC, Shephard A. Patients with sore throat: a survey of self-management and healthcare-seeking behavior in 13 countries worldwide. Pragmatic and observational research. 2020;11:91–102. doi:10.2147/por.S255872

27. Teixeira Rodrigues A, Roque F, Piñeiro-Lamas M, Falcão A, Figueiras A, Herdeiro MT. Effectiveness of an intervention to improve antibiotic-prescribing behaviour in primary care: a controlled, interrupted time-series study. Journal of Antimicrobial Chemotherapy. 2019;74(9):2788–2796. doi:10.1093/jac/dkz244

28. Gulliford MC, Moore MV, Little P, et al. Safety of reduced antibiotic prescribing for self limiting respiratory tract infections in primary care: cohort study using electronic health records. BMJ. 2016;354:i3410. doi:10.1136/bmj.i3410

29. Hoffmann T, Peiris R, Mar CD, Cleo G, Glasziou P. Natural history of uncomplicated urinary tract infection without antibiotics: a systematic review. British Journal of General Practice. 2020;70(699):e714–e722.

30. Merenstein DJ, Barrett B, Ebell MH. Antibiotics not associated with shorter duration or reduced severity of acute lower respiratory tract infection. Journal General Internal Medicine. Apr 15 2024;doi:10.1007/s11606-024-08758-y

31. Driel MLv, Sutter AD, Deveugele M, et al. Are sore throat patients who hope for antibiotics actually asking for pain relief? Annals of Family Medicine. 2006;4(6):494–499. doi:10.1370/afm.609

32. Hoffmann TC, Jones M, Glasziou P, Beller E, Trevena L, Mar CD. A brief shared decision-making intervention for acute respiratory infections on antibiotic dispensing rates in primary care: a cluster randomized trial. Annals Family Medicine. 2022;20(1):35–41. doi:10.1370/afm.2755

33. Hoffmann TC, Légaré F, Simmons MB, et al. Shared decision making: what do clinicians need to know and why should they bother? Medical Journal of Australia. 2014;201(1)

34. Spurling GKP, Del Mar CB, Dooley L, Clark J, Askew DA. Delayed antibiotic prescriptions for respiratory infections. Cochrane Database Syst Rev. 2017;(9)doi:10.1002/14651858.CD004417.pub5

35. Everitt H, Kumar S, Little P. A qualitative study of patients perceptions of acute infective conjunctivitis. British Journal of General Practice. 2003;53(486):36–41.

36. Boaitey KP, Hoffmann T, Baillie E, Bakhit M. Exploring general practitioners’ perception of the value of natural history information and their awareness and use of guidelines’ resources to support antibiotic prescribing for self-limiting infections: a qualitative study in Australian general practice. Australian Journal of Primary Health. 2023;29(6):558–565.

37. Boaitey KP, Bakhit M, Krzyzaniak N, Hoffmann TC. Information about the natural history of acute infections commonly seen in primary care: a systematic review of clinical practice guidelines. BMC Infect Dis. Dec 1 2022;22(1):897. doi:10.1186/s12879-022-07887-1

38. Moles RJ, Stehlik P. Pharmacy Practice in Australia. Canadian Journal of Hospital Pharmacy. 2015;68(5):418–26. doi:10.4212/cjhp.v68i5.1492

39. Australian Bureau of Statistics. 2016 Census All persons quickstats. 22 April, 2024. Accessed April, 22, 2024. https://www.abs.gov.au/census/find-census-data/quickstats/2016

40. Boaitey KP, Bakhit M, Hoffmann T. Mapping the evidence about the natural history of acute infections commonly seen in primary care and managed with antibiotics: a scoping review. BMC Infect Dis. Accepted 17.06.2024

41. De Sutter AIM, Eriksson L, van Driel ML. Oral antihistamine□decongestant□analgesic combinations for the common cold. Cochrane Database Syst Rev. 2022;(1)doi:10.1002/14651858.CD004976.pub4

42. Sheikh A, Hurwitz B, van Schayck CP, McLean S, Nurmatov U. Antibiotics versus placebo for acute bacterial conjunctivitis. Cochrane Database Syst Rev. 2012;(9)doi:10.1002/14651858.CD001211.pub3

43. Spinks A, Glasziou PP, Del Mar CB. Antibiotics for treatment of sore throat in children and adults. Cochrane Database Syst Rev. Dec 9 2021;12(11):CD000023. doi:10.1002/14651858.CD000023.pub5

44. Thompson M, Vodicka TA, Blair PS, Buckley DI, Heneghan C, Hay AD. Duration of symptoms of respiratory tract infections in children: systematic review. BMJ. 2013;347:f7027.

45. Zalmanovici Trestioreanu A, Yaphe J. Intranasal steroids for acute sinusitis. Cochrane Database Syst Rev. 2013;(12)doi:10.1002/14651858.CD005149.pub4

